# The Defense of Shangri-La: A Thought Experiment of Periodic Community-wide Screening in the Future Pandemic

**DOI:** 10.1101/2023.01.03.23284127

**Authors:** Anqi Duan, Jian Li, Zhen Yang, Yungang He

## Abstract

**Objectives:** In a dangerous future pandemic without effective vaccines and medicines, a reliable screening-and-isolation strategy can be the last opportunity to keep critical facilities and communities running and avoid a complete shutdown.

**Methods:** In this study, we introduced an epidemiological model that included essential parameters of infection transmission and screening. With varying parameters, we studied the dynamics of viral infection in the semi-isolated communities.

**Results:** In the scenario with a periodic infection screening once per 3 days and a viral basic reproduction number 3.0, more than 85% of the infection waves have a duration less than 7 days and the infection size in each of the waves is generally less than 4 individuals when the efficiency of infection discovery is 0.9 in the screening. When the period of screening was elongated to once per 7 days, the cases of infection dramatically increased to 5 folds of that mentioned previously. Further, with a weak discovery efficiency of 0.7 and the aforementioned low screening frequency, the spread of infection would be out of control.

**Conclusions:** Our study suggests that frequent periodic screening is capable of controlling a future epidemic in a semi-isolated community without vaccines and medicines.

## 1. Introduction

The history of human civilization is a history of humans struggling against infectious diseases. In 1997, the book “Guns, Germs and Steel: The Fate of Human Society” introduced the influences of pathogens in the development of human beings, and clarified that the emergence of epidemics was closely related to the development of human agriculture, domestication of animals, and population density(Diamond, 1999). Smallpox(Hochman and Souza, 2022), tuberculosis(Rajwani et al., 2022), monkeypox(Bunge et al., 2022), and the sudden emergence of COVID-19(Coronavirus Disease 2019)(Mohamadian et al., 2021) in 2019 all posed great threats to the economy and life of our society because of their significant mortality and infectivity.

In outbreaks of new infectious diseases, in addition to developing vaccines and medicines as quickly as possible, intra-regional interventions are particularly important, including border control, cancellation of large-scale events, and quarantine of infected subjects(Benke et al., 2020; Fong et al., 2020; Ryu et al., 2020; Xiao et al., 2020). These interventions have been shown to be effective in reducing the peak size of epidemics and delaying the spread of diseases, and some researchers have demonstrated that early implementation of rigorous interventions can significantly reduce the size of epidemics(Fan et al., 2021).

Mathematical models can be used to analyze the dynamics of infectious diseases and provide some helpful knowledge on epidemic prevention and control. For example, the SIR (<u>S</u>usceptible-Infectious-<u>R</u>ecovered) model, proposed by Kermack and McKendrick in 1927, was first used to study the black death epidemic in London(Kermack, 1927). Subsequently, Lekone introduced the exposed person into the SIR model, and the expanded model was called the SEIR(<u>S</u>usceptible-<u>E</u>xposed-Infectious-<u>R</u>ecovered) model(Lekone and Finkenstädt, 2006). In addition, it was concluded that the stability of the SEIR model is better than that of SIR after validation by applying the Lyapunov function(Abta et al., 2012). In 2020, Chen introduced population migration into the SEIR model and verified that the prohibition of migration did slow down the spread of disease(Chen et al., 2020).

Due to the profound impacts of infectious diseases on human civilization, many fictions described the images of global disasters caused by infectious diseases. In these fictions, there is often an ideal community as humanity’s last paradise. Great science fictions often have deep glimpses of the future, and many of the old predictions have come true. It is worth discussing, through a thought experiment with profound mathematical models, how to effectively protect a human community in a global pandemic with rigorous scientific theories. Especially when vaccines and medicines are unsuccessful, it is important to formulate public health policies to better protect the community with limited resources. In the present study, we developed a mathematical model to evaluate the effectiveness of the screening-isolation strategy for epidemic control. This study showed that the screening-isolation strategy works well in a semi-isolated community with some specific criteria.

## 2. Methods

In this study, we aimed to evaluate whether a community-wide screening is sufficient to limit contagious diseases in the community when medications and vaccinations are unavailable. It is straightforward to propose a compartmental model for the spread of infectious disease in a semi-isolated community (Fig. 1 A). All members of the community are assumed to be susceptible to the disease that is widely spread in the outside world. To reduce the influx of pathogens, the members rarely move in or out. As the infection is rare in the community, the size of the susceptible population is nearly constant. The aforementioned model can be further simplified to fit a screening-isolation strategy (Fig. 1 B).

**Fig. 1.**
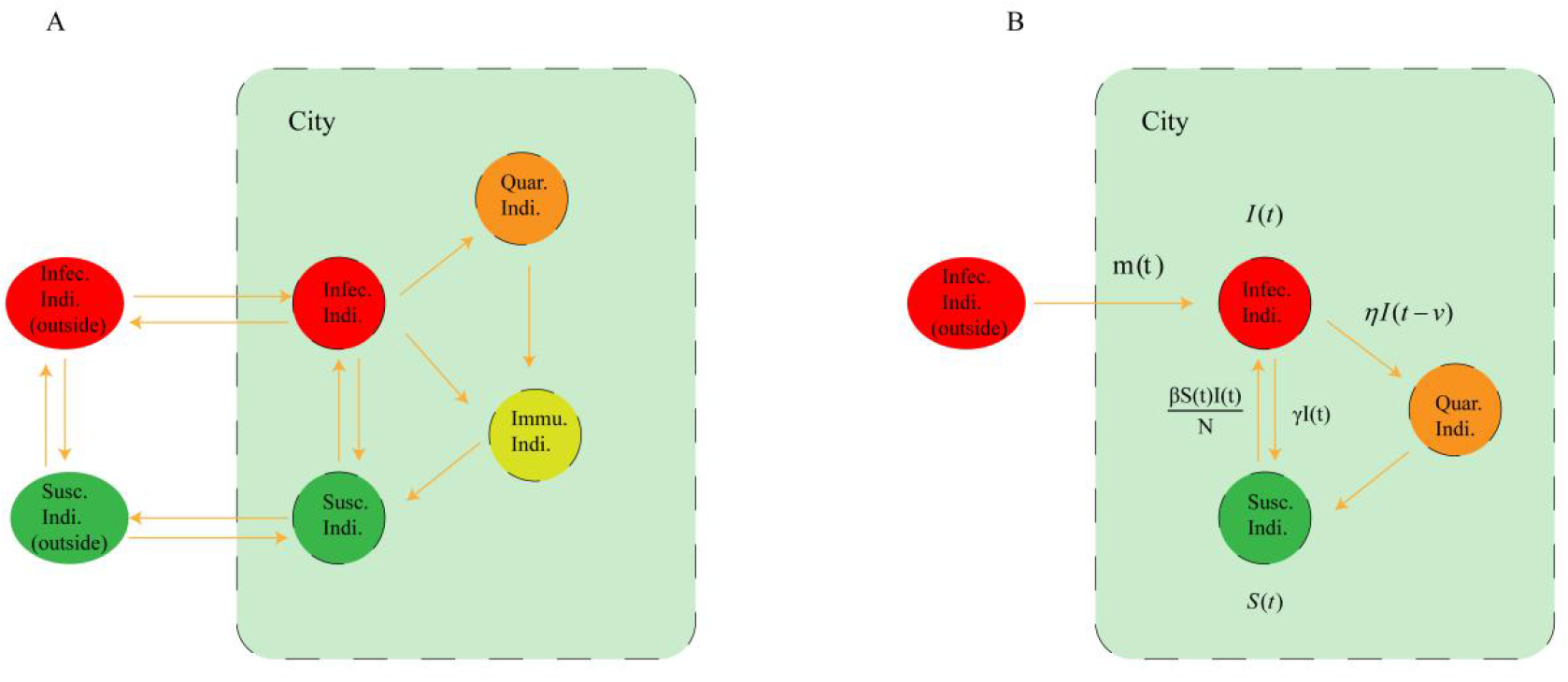
Compartmental models of semi-isolated communities. A. The regular model with movements of individuals. B. The simplified model for the community with a border control.

### 2.1 Modeling

In the regular SIR model, the dynamics of infection was presented by the system of equations below,

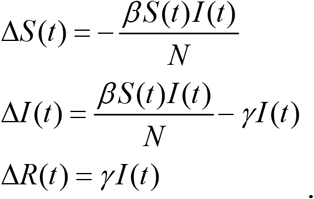

Given the constant population size N, the size of the susceptible group at time t is denoted as S(t) when I(t) is the size of the infected group. ΔS(t), ΔI(t), and ΔR(t) indicate the size changes of the susceptible group, infected group, and recovered group, respectively. The coefficient β denotes the contagious capacity and γ indicates the recovery/death capacity of infected individuals.

In our simplified model (Fig. 1 B), the size change of the contagious group (ΔI) was further divided into the size increase (ΔG) and the size decrease (ΔC). The increase at time t is mainly due to local infection and the influx of infection *m*(*t*) from outside.

Therefore, the size increase can be represented as Δ*G*(*t*) = *βS* (*t*)*I* (*t*) / *N* + *m*(*t*). The decreasing size is caused by the screening efficiency η and recovery/death γ to infected individuals at time t when identified carriers are isolated immediately. Hence, we have the size decrease as Δ*C*(*t*) = *γI* (*t*)+*ηI* (*t* − *v*). There is a time lag ν ≥ 0 if the isolation of infectious individuals happens after the infection. When the infection is rare in the community, the size change of the contagious group was available as Δ*I* (*t*) = *βI* (*t*) + *m*(*t*) −[*γI* (*t*)+*ηI* (*t* − *v*)]. We performed a theoretical analysis of the model to address the time-sensitivity of screening in disease control.

### 2.2 Computer simulation

We investigated the dynamics of infection not only in the aforementioned theoretical analysis but also in computer simulations. The computer simulations presented randomness of infection in different scenarios. In the simulations, the daily increments of infection and influx were assumed to follow a Poisson distribution Δ*G* ∼ *Pois* (*βI* (*t*) + *m*)while the daily decrease of infection was presented as Δ*C* ∼ *Pois* ((*γ*+*η*(*t*))*I* (*t*)). In our simulations, we explored infection dynamics with 3 different levels of daily influx *m* ∈{0.01, 0.1,1}. The different daily screening-isolation efficiency was given as *η*(*t*) ∈{0, 0.7, 0.9}. More specifically, we have *η*(*t*) = 0 if and only if the screening was lacking at the day t. We also assigned an empirical daily recovery/death rate as γ = 0.3 to the simulations. With the given γ, the expected duration of the disease course is about 7.47 days that was obtained in *E*(*T*) = (1 −*γ*)[1 − log(1 −*γ*)] / log 2(1 −*γ*). When the basic reproduction number of the disease (R_0_) indicates the expected number of new infections from a single pathogen carrier, we obtained the daily contagious capacity *β*≈ *R*_0_ / *E*(*T*) in the simulations. The contagious capacities that correspond to different R_0_ were presented in Table 1.

**Table 1.** Basic reproduction numbers of disease with their corresponding contagious capacity.

| $R_0$ | Beta($\beta$) |
| --- | --- |
| 3 | 0.4019 |
| 6 | 0.8038 |
| 8 | 1.0717 |

## 3. Result

### 3.1 Time sensitivity of screening in disease control

It was assumed that the size of the infectious population is capable to expand rapidly in a future pandemic, i.e *βI* (*t*) + *m*(*t*) −*γI* (*t*) ≫ 0. To a successful disease control, it is necessary to reduce the spread of the disease by applying the screening-isolation strategy. In a successful control, the change of the infectious population can be shown as *βI* (*t*) + *m*(*t*) −*γI* (*t*) −*ηI* (*t* − *v*) < 0. Further, when the external influx *m*(*t*) is small, the condition of a successful control can be represented as *ηI* (*t* − *v*) > (*β*−*γ*)*I* (*t*). Without active control before the time t, the size of infection at time t will be far larger than that of before, i.e. *I* (*t*) ≫ *I* (*t* − *v*). If the time lag v is large, it is difficult to meet the aforementioned condition *ηI* (*t* − *v*) > (*β*−*γ*)*I* (*t*). In other words, the spread of the disease is out of control when the time lag is large.

We studied the relationship between the time lag and other critical factors. As the size of infection increases exponentially in the early stage of an epidemic, the infection size at moment t can be approximated as *I* (*t*) = *e*^(*β*−*γ*)*t*^. To ensure that the epidemic is under control, there should be more infectious individuals being isolated than the increasing infection at the moment t, i.e. *ηe*^(*β*−*γ*)(*t* −*v*)^ > (*β*−*γ*)*e*^(*β*−*γ*)*t*^. In this circumstance, we have a constraint for the time lag *v* < [log(*η*) − log(*β*−*γ*)] / (*β*−*γ*). The sensitivity of the time lag to other factors in the screening-isolation strategy was presented in Fig. 2. The spread of disease could be under control only if the parameters fall into the small ‘green’ zones. It is clear that the improvement of screening efficiency η has only a weak contribution to the robustness of screening in disease control.

**Fig. 2.**
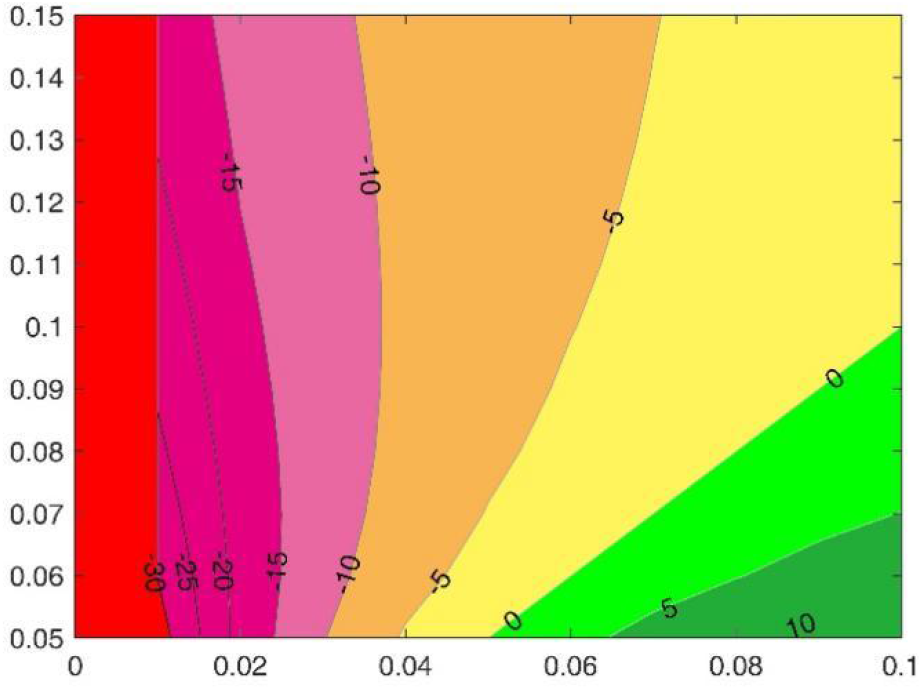
The time sensitivity of screening to transmission parameters. The X-axis indicates the screening efficiency, i.e. *x* = η the y-axis shows the difference between contagious and recovery/death capacities *y* = β-γ the contour lines presents the constraint *c* = [log(*η*) − log(*β*−*γ*)] / (*β*−*γ*). The colorful zones denote the various time sensitivities to the transmission parameters in the disease control. In a successful control, the screening time lag must be smaller than the corresponding constrain.

### 3.2 Effectiveness of the screening-isolation strategy

To verify the screening-isolation strategy in epidemic control, the spread of disease is simulated in the aforementioned model for a semi-isolated community. In the sense of a global pandemic, we assumed the community maintained very limited communication with the outside populations. The probability that the community members contact an infected stranger is assigned to be 10% per day (m=0.1). The recovery/death rate of infected individuals is given as γ = 0.3.

In the scenario with a basic reproduction rate R_0_=3.0, which is close to that of the SARS disease in 2003, the infectious disease was out of control in less than 100 days in all 10 different simulations (Fig 3. A). Alternatively, in the presence of a daily screening-isolation action with an efficiency η = 0.9, the size of infection is well controlled in the community (Fig 3. B). There were no more than 10 infections in a single infection wave in all the simulations, and the average duration of a single wave is only 1.64 days with a standard deviation of ±0.96. The result supports that the screening-isolation strategy is an effective solution for the epidemic control in a semi-isolated community in the absence of medication and vaccination.

**Fig. 3.**
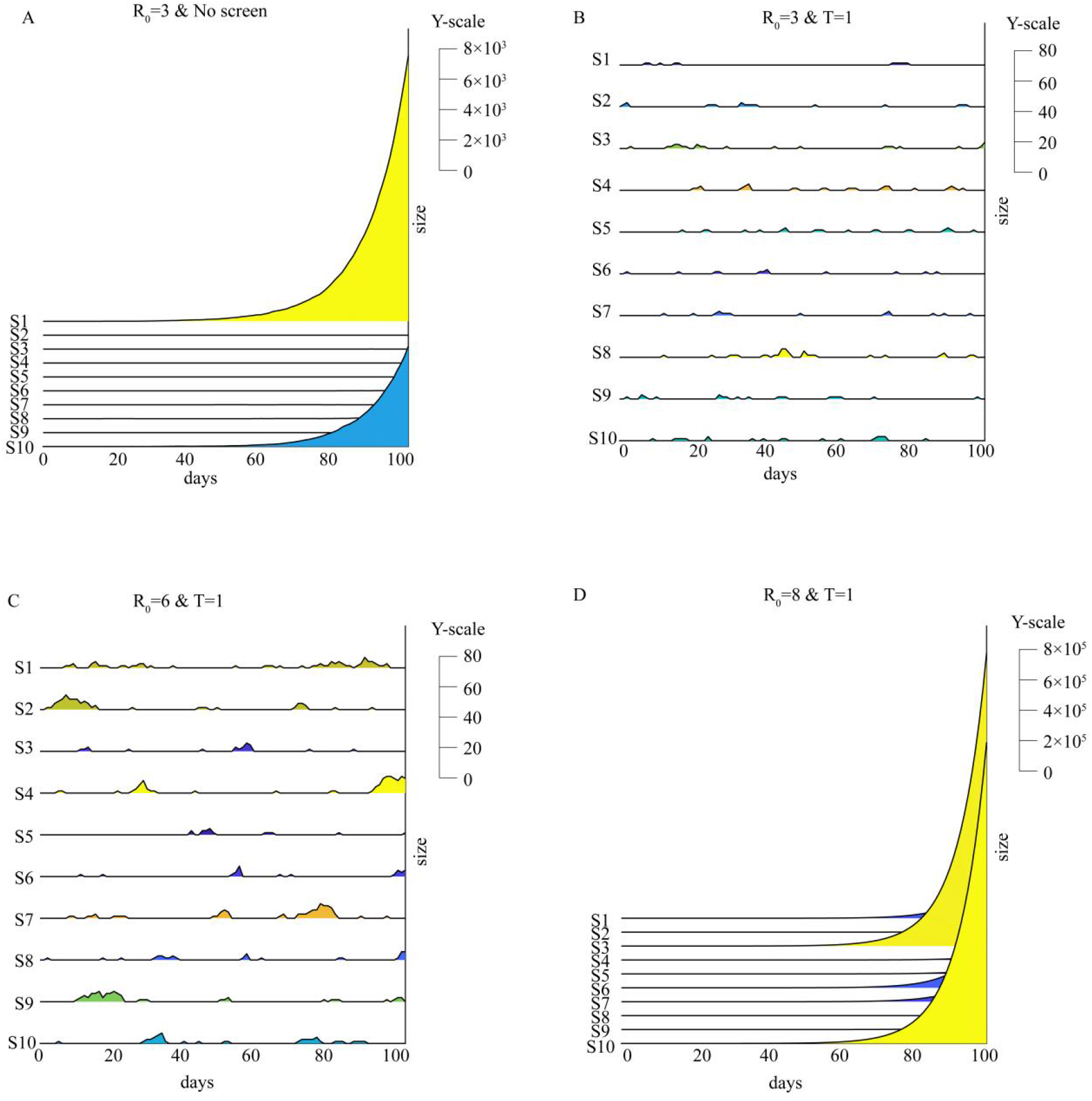
Effectiveness of the screening-isolation strategy in disease control. A. Rapid spreads of infection in 10 simulations with R_0_ 3.0 in the scenario without the periodic infection screening; B. Weak infection waves in 10 simulations with R_0_ 3.0 and the daily screening; C. Significant infection waves in 10 simulations with R_0_ 6.0 and the daily screening; D. The transmission is out of control in 10 simulations with R_0_ 8.0 in the presence of the daily screening.

We further simulated the dynamics of infection in a scenario with R_0_=6.0 (Fig 3. C). In all 10 simulations, total of 576 infections were observed. The infection size was about 4 times that of the aforementioned simulations with R_0_=3.0. Our results showed the screening-isolation strategy still worked well, even if the pre-assigned R_0_ was as dangerous as that of smallpox. However, the screening-isolation strategy failed when the reproduction rate increased to a higher level, says R_0_=8.0 (Fig 3. D). The results suggested that more actions were necessary for disease control in such circumstances with an extraordinary reproduction rate. The screening-isolation strategy can work with other actions that social distancing and mask usage are capable of effectively reducing viral reproduction.

### 3.3 Significance of the screening frequency and efficiency

To evaluate the robustness of disease control, we simulated the dynamics of infection 30 times in our model with a reproduction rate R_0_=3.0 together with different screening frequencies and efficiencies. The results suggested the spread of infection could be under control if the screening was periodically carried out in a frequency of once per 3 days (Fig 4. A & B). When minor infection waves were observed in the 365-day timelines, we were able to characterize the infection waves in the different scenarios (Table 2). In the scenario with a screening-isolation efficiency η = 0.7, the average duration of each infection wave is 3.97 days with std. 4.94 days. The average duration slightly decreases to 3.62 ± 3.88 days (mean ± std.) as the efficiency increases to η=0.9. Although the average durations are comparable, there was a significant difference between infection sizes with the different screening-isolation efficiencies (p-values ≤ 0.002). The average sizes of cumulative infections are 267.46 ±213.83 cases per year and 142.76 ±53.94 cases per year for an efficiency of 0.7 and 0.9, respectively.

**Fig. 4.**
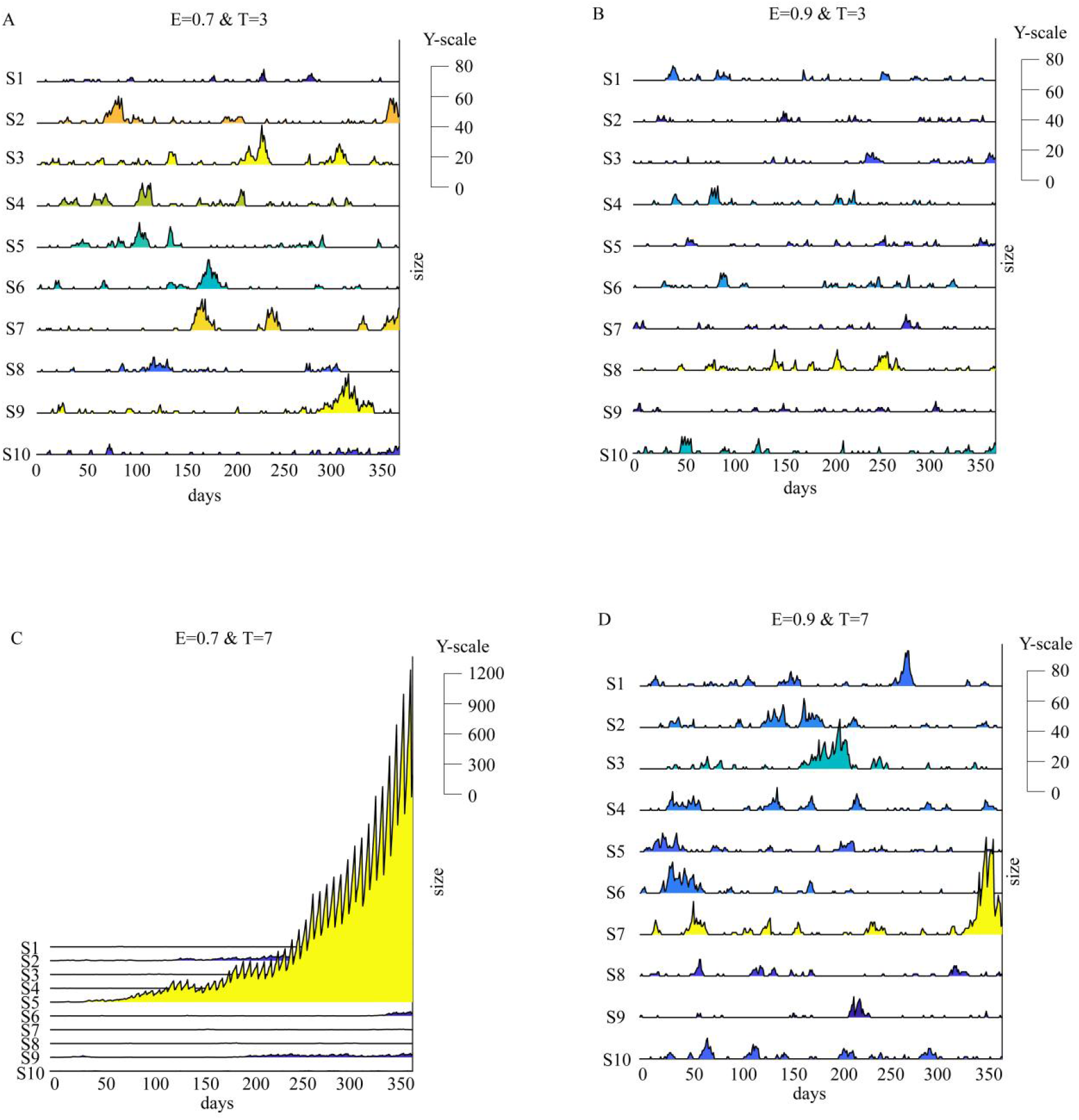
Dynamics of Infection with different screening frequencies and efficiencies. A. Significant infection waves in 10 simulations with the screening efficiency 0.7 and the frequency of once per 3 days; B. Weak infection waves in 10 simulations with the improved screening efficiency of 0.9 and the frequency of once per 3 days; C. The transmission is out of control in 10 simulations with the weak screening efficiency of 0.7 and the reduced frequency of once per 7 days; D. Significant infection waves being under control in 10 simulations with the improved screening efficiency 0.9 and the reduced frequency of once per 7 days.

**Table 2.** Summary of transmission characteristics with different screening frequencies and efficiencies.

|  | Period = 3 days |  | Period = 7 days |  |
| --- | --- | --- | --- | --- |
|  | Efficiency = | Efficiency = | Efficiency = | Efficiency = |
|  | 0.7 | 0.9 | 0.7 | 0.9 |
| Times of waves<br>(in 30 years) | 754 | 797 | 672 | 672 |
| Total duration<br>(days in 30 years) | 2994 | 2884 | 6478 | 3790 |
| Average duration<br>(days ± std) | 3.97<br>($\pm 4.94$) | 3.62<br>($\pm 3.88$) | 9.64<br>($\pm 30.44$) | 5.64<br>($\pm 9.87$) |
| Duration >7 days<br>(in 30 years) | 107 | 98 | 138 | 128 |
| Duration >14 days<br>(in 30 years) | 33 | 24 | 74 | 53 |
| Duration > 30 days<br>(in 30 years) | 5 | 0 | 38 | 15 |
| Total infections per<br>year<br>(± std) | 267.46<br>($\pm 213.83$) | 142.76<br>($\pm 53.94$) | 3.61e4<br>($\pm 9.82e4$) | 575.43<br>($\pm 587.76$) |
| >1000 infections<br>(in 30 years) | 1 | 0 | 18 | 4 |

We further simulated the spread of infection in the semi-isolated community where the disease screening was periodically conducted only once a week (Fig 4. C & D). Our results showed the disease control in the present scenarios was much worse than that of the aforementioned simulations. The cumulative size of infection is as many as 3.61×10^4^ ± 9.82×10^4^ cases per year with a screening-isolation efficiency η=0.7. Alternatively, with the efficiency η=0.9, the size of infection is only 575.43±587.76 cases per year. The average duration of infection waves is 9.64 ± 30.44 days and 5.64 ± 9.87 with an efficiency of 0.7 and 0.9, respectively. More simulations were carried out in scenarios with disease screening once per 2 weeks. In these later simulations, the spread of disease is out of control no matter the efficiency η=0.7 or η=0.9 (Supplementary Figure 1).

### 3.4 Necessity of strict border control

To understand the necessity of strict border control, we simulated the influences of different scales of influx. When the screening was carried out once per 3 days, cumulative cases of infection increased with the increase of both time and influx. As the disease influx is 0.01 cases per day, the average infection is only 14.80±13.94 cases per year (Fig. 5 A). When the influx increase to 0.1 cases per day, the size of infection is about 10-fold of the previous observation with an average of 156.83±47.20 cases per year(Fig. 5 B). With the influx of m=1 cases per day, the annual infection size rises to 2024.83±286.86 cases per year. Our results showed the increase in infection sizes is proportional to the scale of the influx in the scenarios with frequent disease screening. Therefore, strict border control is necessary for successful disease control.

**Fig. 5.**
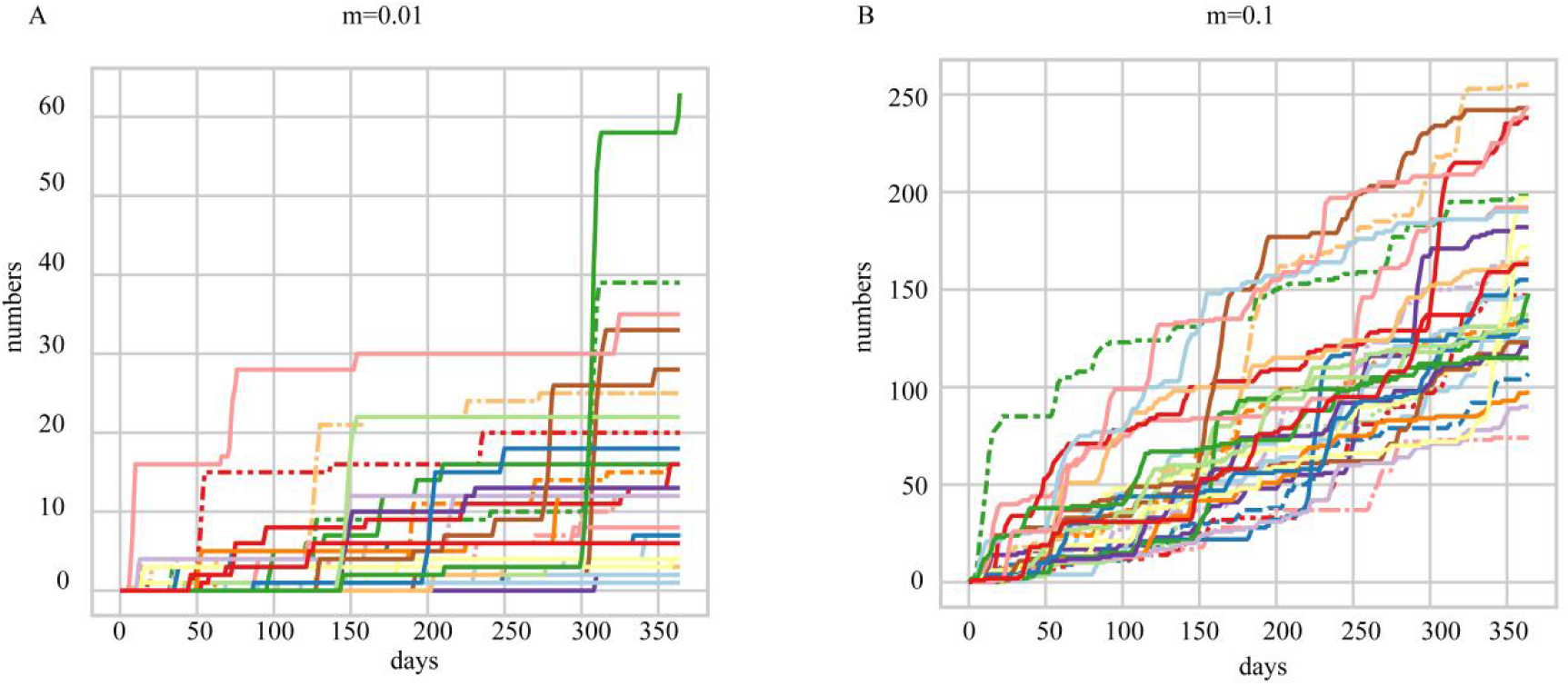
Dynamic of cumulative infection size under different border control. A. The slow increase of cumulative infection in the scenario with a rare influx of infection m=0.01; B. The significant increase of cumulative infection in the scenario with an influx of infection m=0.1.

## 4. Discussion

The present study investigated whether the screening-isolation strategy can protect a semi-isolated community from an untreatable infectious disease in a future pandemic. Our results showed that the screening-isolation strategy works well in general scenarios. This study, usually called “a thought experiment”, is laid out for the purpose of thinking through the consequences of the screening-isolation strategy. Although the present study was based on a simplified model, it is convenient to extend our discussions to more general circumstances.

Traditional screening methods may not work well in future screening. There are multiple high-throughput methods being the common usage in disease screening. Among them, fever screening is one of the most popular methods of epidemic control. It is widely carried out at airport entries and exits to prevent the spread of diseases due to infected travelers. From July 2003–June 2004, fever screening in Taiwan airports identified 40 confirmed dengue cases(Shu et al., 2005). It stopped further transmission caused by the identified cases. The fever screening also played an important role in successfully controlling the epidemic of the severe acute respiratory syndrome (SARS) in 2003(Chiang et al., 2008; Hay et al., 2004). However, the screening does not work in control of all the different diseases. For example, it was suggested that fever screening at Narita International Airport Japan has litter effect on identifying the viral carriers during the influenza (H1N1-2009) pandemic(Nishiura and Kamiya, 2011). In addition, the efficiencies of different screening approach greatly vary for different diseases. Besides its success against SARS and dengue, fever screening has a sensitivity varied from 4.0% to 89.6% and specificity from 75.4% to 99.6%(Bitar et al., 2009).

PCR-based tests are good candidates for future techniques of massive screening during the pandemic. During the pandemic of COVID-19, some screening methods are considered to be more reliable than traditional approaches, such as rapid antigen test(Mertens et al., 2020), antibody test(Cassaniti et al., 2020), and RT-PCR test(Xiang et al., 2020, p. 19). In recent years, biotechnological industries are capable to deliver reagents for antigen tests and RT-PCR tests in several weeks. The antigen tests are designed to directly detect proteins (antigens) in biospecimens. For SARS-Cov-2, the overall sensitivity of the rapid antigen test was around 65.3% while the specificity was 99.9% (95% CI 99.5-100.0%). In asymptomatic individuals, the sensitivity drops as low as 44.0%(Jegerlehner et al., 2021). The PCR-based test is usually more sensitive than other approaches. In a meta-analysis of 23 studies of SARS-Cov-2, using nasopharyngeal swabs as the gold standard, the pooled sensitivities were 86 percent for nasal swabs, 85 percent for saliva samples, and 68 percent for throat swabs. The combination of nasal and throat swabs may be as high as 97% (95% CI 93-100%)(Tsang et al., 2021). The excellent performance of PCR-based tests makes it a good candidate approach for community-wide screening in a future pandemic.

In the present study, we assumed that the infectious and detectable periods span the same time interval during the disease progression. However, with the time delay parameter (v) mentioned above, it is convenient to extend our work to circumstances where the infectious and detectable periods cover different time intervals. If the infection is detectable before the carriers are capable to transmit the pathogen to others, we can represent the epidemic dynamics in the aforementioned model with an ignorable time lag v=0. Consequently, disease transmission would be more likely to be eradicated in the community. Alternatively, if the infection is detectable after the onset of the transmission period, the dynamic of the epidemic can be shown in our model with a significant time lag v>>0. In situations with significant time lags, disease control can be a daunting task. Our thought experiments supplied a glimpse into the disease control of a semi-isolated community in a future pandemic. The results suggested that periodic community-wide screening is capable to control the spread of serious diseases in the community without the intervention of medication and vaccination.

Although labor and financial costs may not be a big issue in disease control, it is still a challenge how to reduce their interference with the daily operation of the community. Further, the control team may have to deal with community residents’ physiological problems, such as PTSD, etc. It is necessary to carry out more thought experiments in different fields to supply a full solution to the future pandemic.

## 5. Conclusions

Overall, our study suggests that regular and periodic screening for epidemics in semi-isolated communities without vaccines and medications can be a good way to control epidemic transmission. More notably, the frequency and efficiency of screening and the migration rate have an impact on the size of the infection.

## Data Availability

All codes and other material used in this study are publicly available upon request

## Abbreviations

SIR: <u>S</u>usceptible-Infectious-<u>R</u>ecovered
SEIR: <u>S</u>usceptible-<u>E</u>xposed-<u>I</u>nfectious-<u>R</u>ecovered
COVID-19: Coronavirus Disease 2019

## Author contribution

Y.H. and Z.Y. conceptualized the study; Y.H. and A.D. developed the methodolodymethodology; A.D. and J.L. performed the investigation; Y.H. and Z.Y. supervised the project; Y.H., A.D. prepared the manuscript.

## Acknowledgements

Not applicable.

## Appendix A. Supplementary data

Supplementary Figure 1

## Data availability

All codes and other material used in this study are publicly available upon request.

