## Supplementary Figure 1 for "The Defense of Shangri-La: A Thought Experiment of Periodic Community-wide Screening in the Future Pandemic"

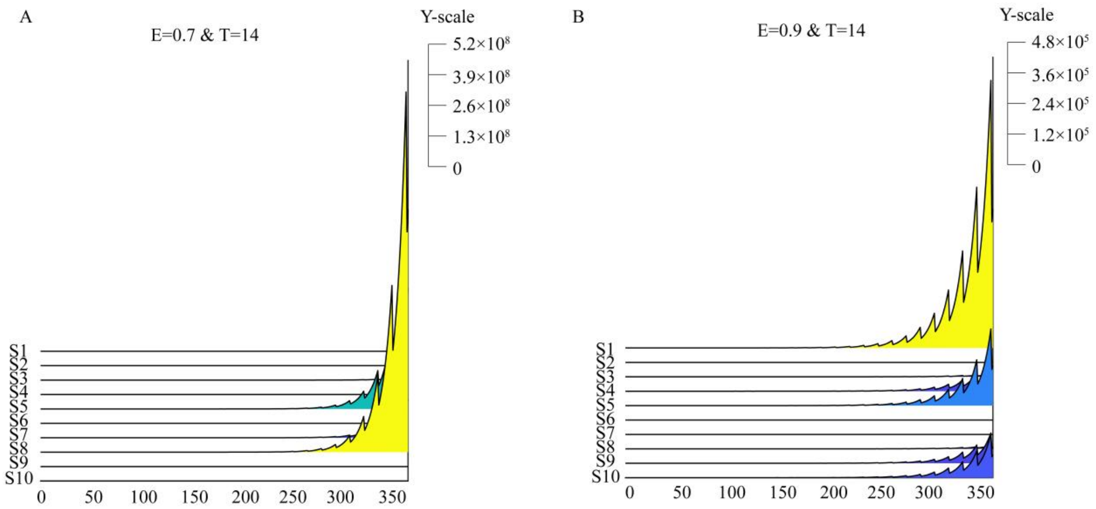


**Sup Fig. 1 Infection dynamics at different efficiencies when R0=8 and screening frequency is two weeks.** A. At a screening efficiency of 0.7, transmission is completely out of control; B. At a screening efficiency of 0.9, the infection situation is equally unobjectionable.
